# SHIVIR - An Agent-Based Model to assess the transmission of COVID-19 in India

**DOI:** 10.1101/2022.05.26.22275624

**Authors:** M.S. Narassima, Denny John, S.P. Anbuudayasankar, Guru Rajesh Jammy, Rashmi Pant, Lincoln Choudhury

**Affiliations:** Academics, Great Lakes Institute of Management, Chennai, Tamil Nadu, India; Department of Public Health, Ramaiah University of Applied Sciences, Bengaluru, Karnataka, India; Department of Mechanical Engineering, Amrita School of Engineering, Coimbatore, Tamil Nadu, India; Health Nutrition and Population, The World Bank, New Delhi, Delhi, India; Health Research, Society for Health Allied Research and Education India, Gurgaon, Haryana, India; DLF capital Green, Moti Nagar, New Delhi, Delhi, India

**Keywords:** Agent-Based Modelling, COVID-19, Disease Modelling, India, Infectious diseases

## Abstract

**Background:** COVID-19 has tormented the global health and economy like no other event in the recent past. Researchers and policymakers have been working strenuously to end the pandemic completely.

**Methodology/Principal Findings:** Infectious disease dynamics could be well-explained at an individual level with established contact networks and disease models that represent the behaviour of the infection. Hence, an Agent-Based Model, SHIVIR (Susceptible, Infected, Admitted, ICU, Ventilator, Recovered, Immune) that can assess the transmission dynamics of COVID-19 and the effects of Non-Pharmaceutical Interventions (NPI) was developed. Two models were developed using to test the synthetic populations of Rangareddy, a district in Telangana state, and the state itself respectively. NPI such as lockdowns, masks, and social distancing along with the effect of post-recovery immunity were tested across scenarios.

The actual and forecast curves were plotted till the unlock phase began in India. The Mean Absolute Percentage Error of scenario MD100I180 was 6.41 percent while those of 3 other scenarios were around 10 percent each. Since the model anticipated lifting of lockdowns that would increase the contact rate proportionately, the forecasts exceeded the actual estimates. Some possible reasons for the difference are discussed.

**Conclusions:** Models like SHIVIR that employ a bottom-up Agent-Based Modelling are more suitable to investigate various aspects of infectious diseases owing to their ability to hold details of each individual in the population. Also, the scalability and reproducibility of the model allow modifications to variables, disease model, agent attributes, etc. to provide localized estimates across different places.

**Author Summary:** The world has witnessed several infectious disease outbreaks from time to time. COVID-19 is one such event that tormented the life of mankind. Healthcare practitioners, policymakers, and governments struggled enormously to handle the influx of infections and devise suitable interventions. Agent-Based Models that use the population data could cater to these requirements better. Hence, we developed a disease model that represents various states acquired by COVID-19 infected individuals. The contact network among the individuals in the population was defined based on which the simulation progresses. The effect of various Non-Pharmaceutical Interventions such as lockdowns, the use of masks and social distancing along with post-recovery immunity were enacted considering two case studies viz. population of Rangareddy district and Telangana state. The capability of these models to adapt to different input data fields and types make them handy to be tailored based on available inputs and desired outputs. Simulating them using local population data would fetch useful estimates for policymakers.

## 1. Introduction

Infectious diseases affect the economy, healthcare systems, public health, and society [1]. There have been 1438 epidemics reported by the World Health Organization (WHO) between 2010 to 2018 [2]. Though all these events have created a substantial impact across multiple dimensions, COVID-19 has made a colossal impact since its outbreak in Wuhan, China in December 2019. The pandemic has presently marked its presence across 224 territories causing 442,413,066 infections with 6,001,844 deaths and 375,259,135 recoveries as of March 04, 2022 [3]. Researchers, healthcare fraternity, and governments worldwide have been working in tandem to devise policies to curtail the spread of pandemic [4,5]. These are achievable by employing mathematical models that could assess multiple aspects like the transmission dynamics, effect of Non-Pharmaceutical Interventions (NPI), the capacity of health infrastructure, etc. [6–8].

The capability of modelling and simulation to replicate the behaviour of real-time systems and generate good estimates has attracted researchers from a range of sectors, especially healthcare and engineering [9]. Of the two broad categories of simulation viz. Compartmental and Agent-Based Modeling (ABM), latter can address infectious diseases more precisely owing to its ability to accommodate agent-level details. The bottom-up approach of these models wherein the behaviour of individual agents cumulates to give the overall behaviour of the system makes it more suitable compared to the compartmental models. ABM approach in the past has been employed to plan evacuation strategies for airborne infections [10], devise methods to administer vaccines for influenza [11], prevent the spread of measles [12], tuberculosis [13], and smallpox [14], and others. Proper use of simulations could help the developing countries with limited healthcare resource settings, like India for planning capacity based on estimates. Some researchers have adopted compartmental models such as Susceptible (S), Hospitalized or Quarantined (H), Symptomatic (I), Purely Asymptomatic (P), Exposed (E), Recovered (R) and Deceased (D) (SIPHERD) [15], Susceptible (S), Exposed (E), Infective (I), and Recovered (R) (SEIR) [16], or mathematical models [17] in the context of COVID-19. The advantageous approach i.e., ABMs, has been used in several aspects of COVID-19 such as to safeguard the vulnerable population [18,19], devise NPI such as lockdowns, use of mask, and social distancing [6,19], schedule location and time-dependent contacts [19,20], assess transmission [20], etc.

The model proposed in the study adopts an ABM approach that simulates the given synthetic population to assess the transmission dynamics. The NPI such as lockdowns, use of masks, and social distancing along with the effect of post-recovery immunity have been included to compare the effects of imposing interventions concurrently. It also has the flexibility to account for changes in the values of parameters, contact network, disease model, and NPIs imposed. These would be helpful to assess the transmission of infectious diseases using regional synthetic populations and devise interventions appropriately [21].

## 2. Modelling of infectious diseases

Modelling of infectious diseases requires interactions among the agents/ entities that govern the transmission of these close contact infections [22]. These cannot be incorporated into the compartmental models that work based on the general population model by distributing the population across various compartments [23]. Each compartment corresponds to a health state such as Susceptible, Exposed, Infected, Recovered, etc. Partitioning/ Compartmentalization is based on the required number of states. In these models, the transitions are governed by rates of flow and variables influencing the flow [24]. Heterogeneity, which is needful for infectious diseases is achieved using time-dependent contact mixing patterns, networks across the community, and seasonality [25,26]. Yet, these models might be less accurate as they neither incorporate agent-level details nor involve interactions among agents. Establishing a contact network within the population being studied or agents is essential to achieve the same. This has been proven by researchers in the past where Compartmental and ABM have been compared. The capability of the latter to track each agent during the simulation enables it to deliver additional agent-specific results [27]. This is because the latter models adopt a simulation-based approach in addition to the set of equations that govern the process [27]. The bottom-up approach makes the incorporation of heterogeneity easier by defining attributes at agent level. Unlike the compartmental models where the transition is governed by variables, the behaviour of agents and contact networks govern the transition between the states in ABMs [6,27]. Despite the advantages of ABMs to inculcate more heterogeneity and population dynamics, the implementation is subject to the computational capabilities and technical expertise of the modellers [27,28].

## 3. SHIVIR - Agent-Based Model

The SHIVIR (Susceptible, Infected, Admitted, ICU, Ventilator, Recovered, Immune) model has been developed based on the understanding of the dynamics of COVID-19 among the infected people from risk of infection to hospitalisation and death. Inputs from epidemiologists, public health practitioners, and biostatisticians were useful for the development of the model as presented successively.

### 3.1. Agent Creation

The creation of agents forms the base for ABM studies. In the studies in which the agents represent the actual population of any territory, a synthetic population approach can be employed. People in the population are modelled as agents for these simulations. The population data from Census could be used to generate the synthetic population based on the required fields/ characteristics to be assigned to each agent[29]. Data cleansing is to be done to eliminate the records with invalid or missing information for any of the required fields. Unique attributes could be assigned to each of the agents in the population to closely represent the actual population studied. The agents could be distinguished into various cohorts to assist a sub-group analysis of measured outcomes.

### 3.2. Disease Model

The progression of infection across various phases in an infected human need to be defined by a disease model. The SHIVIR model was developed with the following states:

1. Susceptible/ Healthy: Every agent in the population is assigned this state. Agents (people) in this state are susceptible to infection upon contact with an infected individual.
2. Infected: Agents who have acquired the infection and are in incubation period.
  a. Asymptomatic: The infected agents who do not show any symptoms. They are unnoticed and continue to transmit the infection to others till recovery.
  b. Symptomatic: The infected agents who exhibit symptoms. They are diagnosed and admitted in isolation. They do not spread the infection to others post-admission. These agents recover when they are in any of these three states: Admitted, ICU, and Ventilator.
3. Admitted: Initial stage of treatment for symptomatic agents. Indicates mild or moderate infection level.
4. ICU: Agents with a higher infection level. These agents require additional care and extended treatment.
5. Ventilator: Agents with severe infection levels. They need the support of a ventilator for breathing.
6. Recovered and Immune: Agents who have recovered from the infection. They are assumed to possess immunity for a certain duration post-recovery. They are not susceptible to infection during this period.
7. Deceased: Agents who have failed to recover from the infection and died. They are no more a part of the simulation.

All the created agents are assigned “Healthy” status at the start of the simulation. Every agent can exist in any of the above-mentioned states at an instant. Based on the contacts made by agents during the simulation, they are subject to acquire infection upon transmission from an infected agent. The agents then undergo a series of transitions across the different states. Duration of existence in each state and probability of transition to successive states are governed by the parameters defined in Table I. From the descriptions of the states, it is interpretable that the contact network plays a significant role in transmitting the infection across the population. The Timestep considered is day as it is suitable to define the transition between the states and contact rate. Based on the transitions of agents, the counts of agents in each state described in the disease model would be recorded at the end of each day, for the three age groups considered viz. kids (age < 5), adults (5 ≤ age ≤ 59), and elders (age > 59). The model allows alteration (addition/ removal or modification) of any states and the cohorts considered.

**Table I:** Parameters during lockdown phases

| Lockdown | Duration | Days | Lockdown Stringency (%) | Contact rate (%) | Close contacts (%) |
| --- | --- | --- | --- | --- | --- |
| Phase 1 | 25 March 2020 – 14 April 2020 | 21 | 75 | 42.80 | 90.75 |
| Phase 2 | 15 April 2020 – 3 May 2020 | 19 |  |  |  |
| Phase 3 | 4 May 2020 – 17 May 2020 | 14 |  |  |  |
| Phase 4 | 18 May 2020 – 31 May 2020 | 14 |  |  |  |
| Unlock 1.0 | 1 June 2020 – 30 June 2020 | 30 | 50 | 56.44 | 81.50 |
| Unlock 2.0 | 1 July 2020 – 31 July 2020 | 31 |  |  |  |
| Unlock 3.0 | 1 August 2020 – 31 August 2020 | 13 | 25 | 70.08 | 72.25 |
| **Post Unlock** | After 31 August 2020 | NA | 0 | 100 | 63 |

### 3.3. Contact Network

Contact network was established henceforth based on the characteristics of agents in the population. It is a factor that majorly affects the spread of infection across the population. The present model considered the contacts made at home, schools, and work depending on place and age as defined by Prem et al., (2017). This helped segregate the contacts like those in the closer circle (home) and external. For each agent in the population, a list of close circle contacts was defined based on the closeness of their locations i.e., the probability of two agents being in each other’s closer circle is inversely proportional to the distance between them, as in equation (1). The following two assumptions were made to work this out:

i. Two people who are farthest in the population have zero probability of meeting
ii. The probability of people with the same Geographic Information System (GIS) coordinates to meet is ‘1’, which is certain in any scenario.

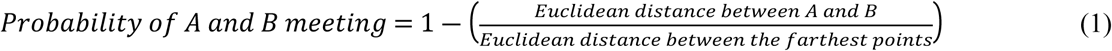

where ‘A’ and ‘B’ are any two people within the population.

The proportion of contacts made with external people was proportionately reduced for stricter lockdowns while the ones in the closer circle were retained. Also, the number of contacts made by each person is to be determined based on the location and age. For this, the results of a study by Supriya Kumar et al., (2018) that determined the contact rates for close-contact infections were utilized. Supriya Kumar et al., (2018) defined the number of contacts made by each person in Ballabgarh district in India, which were used to proportionately calculate the contacts made by people in districts of Telangana. This was accomplished by assuming a density-dependent contact rate i.e., contact rate and population density are directly proportional to each other. The distributions followed by these contact rates were generated using the Input Analyzer tool of Arena (a Rockwell Automation software) that generates distributions followed by the input dataset[32].

## 4. Results of application of the SHIVIR model in Telangana state, India

The disease model was replicated to test the performance of the chosen population of Rangareddy district and Telangana state, India. Initially, the Rangareddy model was created using AnyLogic while that of Telangana was coded using Python. The former is a relatively simpler model that was developed during the initial stages while the latter involves complex dynamics and interventions. To make the mode reproducible, we coded the same using Python.

### 4.1. Timestep

The Timestep chosen for the study is days as it would be meaningful to present these time series estimates daily and also that the contact rates and COVID-19 related variables are defined in days. A healthy individual who has been infected through contact on a day would not acquire a secondary infection. Duration of the existence of each individual in state increments each day to transform to the next state upon reaching the defined time of existence in that state. The code iterates through the entire population every day and governs their contacts with other agents in the population. The state of existence of agents on a particular day governs their behaviour or interaction with other agents.

### 4.2. Synthetic population approach for Rangareddy district and Telangana state

The initial model for Rangareddy was run with 5,48,323 agents representing 10.35 percent of the population of the district as per the 2011 Census of India [33]. The population was divided into three age groups as mentioned in Section 3.2.

The second study on Telangana used data consisting of 31,738,270 people as per the 2011 Census of India, as generated by Sayeed, 2018[29]. Each agent consisted of unique identifiers/ parameters such as:

i. Geocoordinates
ii. District Code
iii. Household ID
iv. Age

Additionally, a unique ID to each agent of the population was mapped for direct reference. Initially, 30 records that did not contain one or more of the required information were discarded and 35,003,674 operable agents were retained. These represented 90.67 percent population of Telangana [33,34]. The Initial health status of all the created agents was set as “Healthy”, indicating their susceptibility to infection.

### 4.3. Establishment of contact network

The number of contacts made by every agent daily and proportionate reduction during the lockdowns were designed as explained in Section 3.3. For the Rangareddy model, different distributions were generated based on the stringency of lockdowns by proportionately reducing the contacts made by every agent per day.

For the Telangana model, the contact distributions were proportionately altered based on the lockdown stringencies such that the proportion of contacts in closer circles increased whilst reducing the overall contact rate. A close contact list was defined for each agent in the population based on the Euclidean distance approach considering the GIS coordinates of the agents. This is to set the list of contacts with which an agent would more likely interact i.e., close contacts with whom there are more chances of interactions during lockdowns. This was done considering the actual reduction in external contacts that happen during lockdowns.

### 4.4. Intervention scenarios

In the Rangareddy model, imposing lockdowns was the only NPI modelled across three different scenarios. The base scenario was a ‘no lockdown’ with no interventions. Two other scenarios represented 50 and 75 percent stringencies imposed. The number of contacts was defined based on the cumulative contacts made at home, office, school, and others based on the results of Prem et al., (2017). To enact the lockdown scenarios, the workplace and other location contacts were reduced proportionately (50 and 75 percent) whilst completing eliminating the contacts at school to indicate closure of schools.

In the Telangana model, the lockdown was imposed as per the Indian scenario [35]. Further, to observe the effects of post-recovery immunity and adoption of mask and distancing, six scenarios were enacted. The scenarios are MD100I90, MD75I90, MD50I90, MD100I180, MD75I180, and MD50I180. An example for the nomenclature of scenarios are as follows:

i. MD100: ‘100’ succeeding ‘MD’ indicates that 100 percent of the population follows the use of masks and social distancing.
ii. I90: ‘90’ succeeding ‘I’ indicates the post-recovery immunity days when a recovered person is not susceptible to infection.

The stringency of lockdowns and the contacts made were during each phase are presented in Table I. Higher the stringency of lockdown, lower the contact rate. Contact rate indicated in the percentage of contacts made as compared to a no lockdown/ normal scenario. It is seen that the proportion of close contacts is higher for more stringent lockdowns indicating higher contacts within the locality or at home.

### 4.5. Data inputs to the model

Following the development of the disease model, establishment of contact network, and agent creation, simulation has to be performed to assess the spread of infection across the given synthetic population. The input variables used for the simulation of models are presented in Table II. For both the districts, the population densities of Rangareddy and other districts were used to derive the contacts made per day considering a population-dependent contact rate [36]. The synthetic population generated forms the basis of simulation as the entire simulation aims to estimate the measures for the input population.

**Table II:**
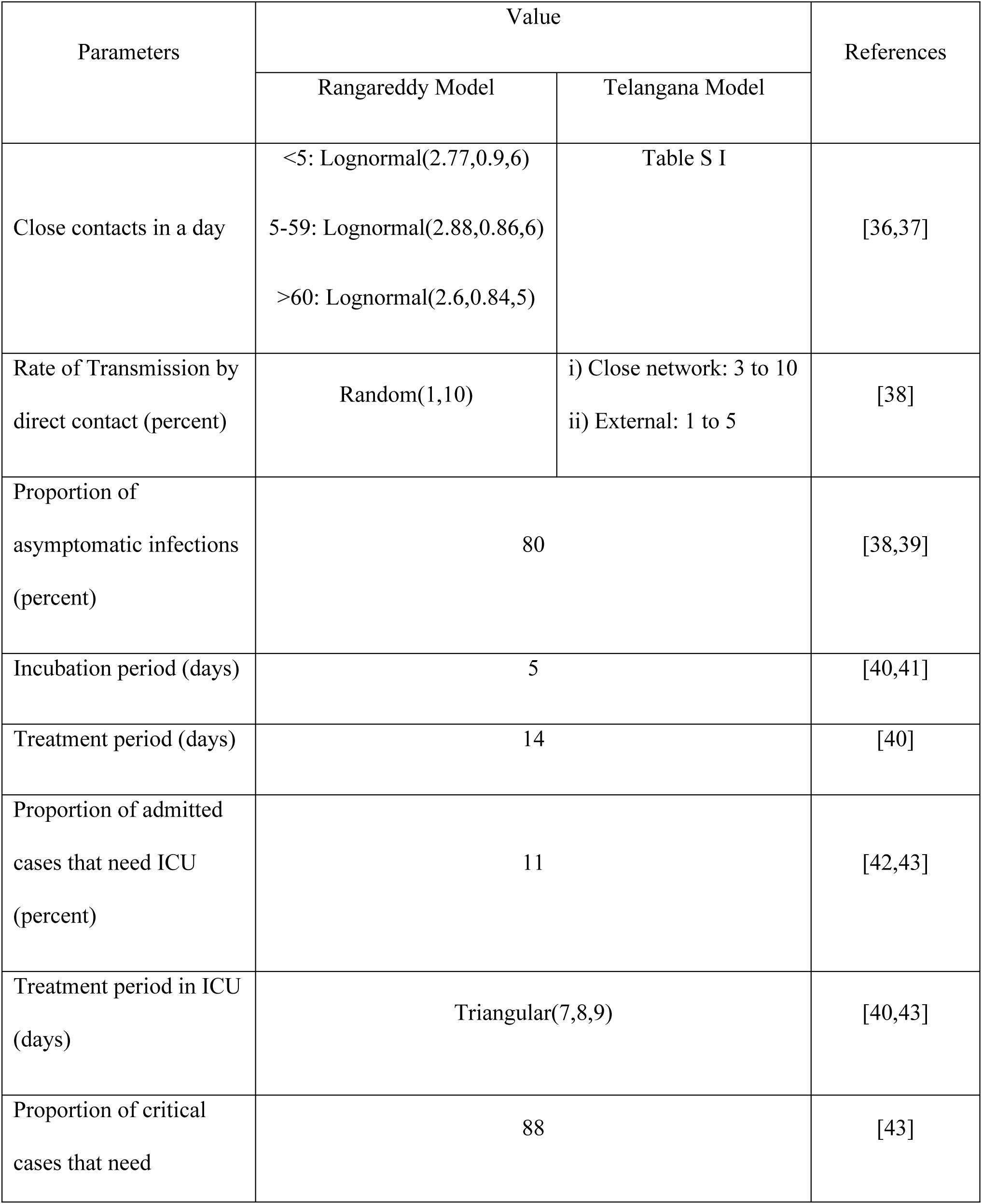
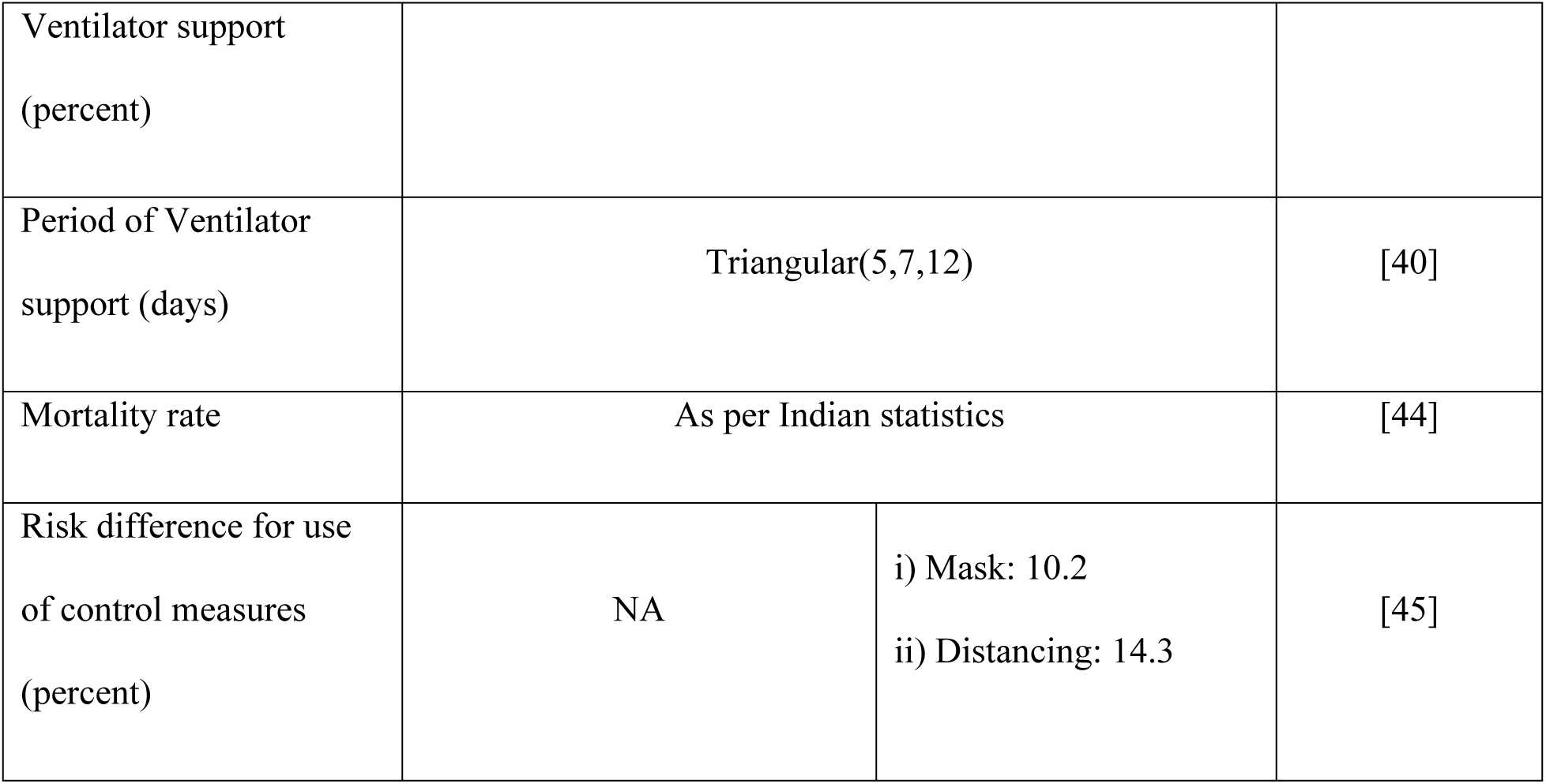
Input variables to the models

**Table III:** Mean Absolute Percentage Error across the scenarios

| Start date | Till date | MD100I90 | MD75I90 | MD50I90 | MD100I180 | MD75I180 | MD50I180 |
| --- | --- | --- | --- | --- | --- | --- | --- |
| 26/04/2020 | 31/05/2020 | 17.35 | 10.46 | 10.93 | 6.41 | 35.23 | 10.50 |
| 26/04/2020 | 30/06/2020 | 78.68 | 43.51 | 20.25 | 20.06 | 112.63 | 18.90 |

## 5. Validation of the results

The model was simulated for 365 days by introducing an infected agent to the population. The code was run on a High-Performance Computing server facility of Amrita Vishwa Vidyapeetham, India. The estimates provided by the model are governed by the variables that were gathered during the initial stages of the pandemic. Also, the lockdown phase considered was based on the initial phases that were imposed in India for 142 days [35]. The model increased the contact rates proportionately for each of the unlock phases and assumed a 100 percent normalcy post 142^nd^ day, which marks the end of Unlock 3.0 (Table I). The symptomatic infections in the model that indicate the diagnosed and identified positive cases were plotted against the actual values for the period from 26 April 2020, the date from which the actual values were available till the end of lockdown phase 4 (31 May 2020) [44]. Mean Absolute Percentage Errors for each of the simulated scenarios were found. The scenario MD100I180 had the least MAPE of 6.41 percent till the end of lockdown phase 4 i.e., 31 May 2020. The scenarios MD75I90, MD50I90, and MD50I180 had MAPE values around 10 percent during this period. Whilst considering the unlock phases (after 31 May, 2020), the error increased. In the simulated model, the stringency of lockdowns was reduced from 75 to 50 percent post phase 4 which is a possible reason for the gap between the actual and forecast to widen.

The actual value is lesser than the forecast indicating the cumulative effects of various interventions apart from lockdowns that offered protection, similar to the Swiss cheese model [46]. Offices that had the capabilities to offer work-from-home continued to the practice. Schools and colleges were not opened as education continued to happen online [47]. Also, there were restrictions imposed on mass gatherings and in public places to prevent transmission on a larger scale [48]. Self-awareness among the public to avoid getting infected is a major driver that flattened the transmission curve [49]. All these measures resulted in overall increased protection on a societal level. At an individual level, avoiding over-crowding, going to closed environments where there are higher chances of infection through suspended particles, wearing face masks, using sanitizers, maintaining social distancing, etc., proved to be effective [45,48].

Of all these interventions and control strategies that could help to curtail the spread of infection, the Telangana model included only lockdowns, use of masks, and social distancing with the effect of post-recovery immunity. The model also considered reducing the proportion of close contacts and external contacts at school, workplace, and others based on the stringency of lockdown. The exclusion of other parameters such as the proportion of the population with work from home capabilities [5], contact tracing to identify and isolate the close contacts and asymptomatic carriers [7,50], and indirect transmission through suspended particles, objects, etc. [20], underreporting of the cases [51], and others, could be some of the reasons for the gap.

Face mask and social distancing reduce the transmission by 10.2 and 14.3 percent respectively [45]. Owing to their importance, the State and Central governments across India have been imposing penalties for not wearing masks [52,53]. Yet, right from the early phases of the pandemic, a significant proportion of the population has not been using face masks. The reported values are 44 percent in India as of September 2020 [54] and May 2021 [55], 52 and 45 percent in Chandigarh [56] and Hyderabad [57] respectively as of January 2022. Discomfort being the highly reported reason among the public is irrelevant considering the objective of wearing masks. Awareness programs need to be held to emphasize the importance of acting socially to protect oneself and others.

## Conclusions

The study presented the framework to use ABM for studying infectious diseases using a synthetic population approach. Infectious disease dynamics are well-explained on an individual level with contact networks than compartmental which is the key idea behind the adoption of this approach [6,58]. The development of a suitable disease model to represent the behaviour of COVID-19 was the initial process. The disease model, SHIVIR was developed in this study to assess the effects of various NPIs and transmission dynamics of COVID-19. The models were developed using AnyLogic (Rangareddy) and Python (Telangana). The former model is less complex as it involved only lockdown as NPI whilst the latter model included the effect of masks, social distancing, and post-recovery immunity additionally. The Telangana model had a more complex contact network that mapped close circle contacts using GIS coordinates. Also, in addition to age, the latter model included GIS, household ID, district code. The simulation was run for 365 days across six different scenarios involving varied combinations of the NPIs. The lockdowns were imposed per the actual ones that were in place in India. The forecast and actual curves matched closely during the initial lockdown phases. After the beginning of the unlock phases, the reduction of lockdown stringency and increase in contact rate in the model spiked the estimates generated by the model. Contrarily, the slope of the actual curve was much lesser than those of the estimates because of the cumulative interventions in real-time.

The code is expandable and reproducible in terms of the synthetic population being used, the attributes mapped to each agent, addition/ deletion/ modification of existing states in the existing (SHIVIR) model, behaviour of agents in each state, transition rules, and probability between the states, duration of stay in each state, the interaction between the agents (contact network), contact rate and probability, NPIs imposed and their associated effects on transmission, level of NPI, etc. These determinants make the model more suitable for different scenarios with varied requirements. ABM models as in this study could help the governments and policymakers to understand the lower-level dynamics better to devise localized NPI strategies that are more suitable for infectious diseases [21]. Also, conservative steps can be taken during such events to anticipate the worst possible outcomes and enhance the preparedness of healthcare systems and resources. Future works could focus on developing similar models using the ABM logic to estimate the transmission dynamics of other infectious diseases. Other aspects such as vaccination pattern, interaction among the agents, contact tracing, etc., could be analyzed in the context of an event. The estimates provided by such models would help to be equipped to handle uninvited events in the future. The addition of more dimensions such as Social Determinants of Health, schedule-based contacts, workplace restrictions, modes of transportation used, etc., might provide more accurate estimates. However, these are subject to data availability and time within which the models need to be acted.

## Data Availability

The synthetic population used, the AnyLogic model along with the input data are available at: https://cloud.anylogic.com/model/7cd10c0c-f1c1-4b8f-9aac-0bf37a45379a?mode=SETTINGS and
https://osf.io/utmhg/?view_only=05ac26fc100645be8b1bba6557d606be.
The code developed using Python is available at: https://doi.org/10.6084/m9.figshare.19121939.

https://cloud.anylogic.com/model/7cd10c0c-f1c1-4b8f-9aac-0bf37a45379a?mode=SETTINGS

https://osf.io/utmhg/?view_only=05ac26fc100645be8b1bba6557d606be

https://doi.org/10.6084/m9.figshare.19121939

## Author Contributions

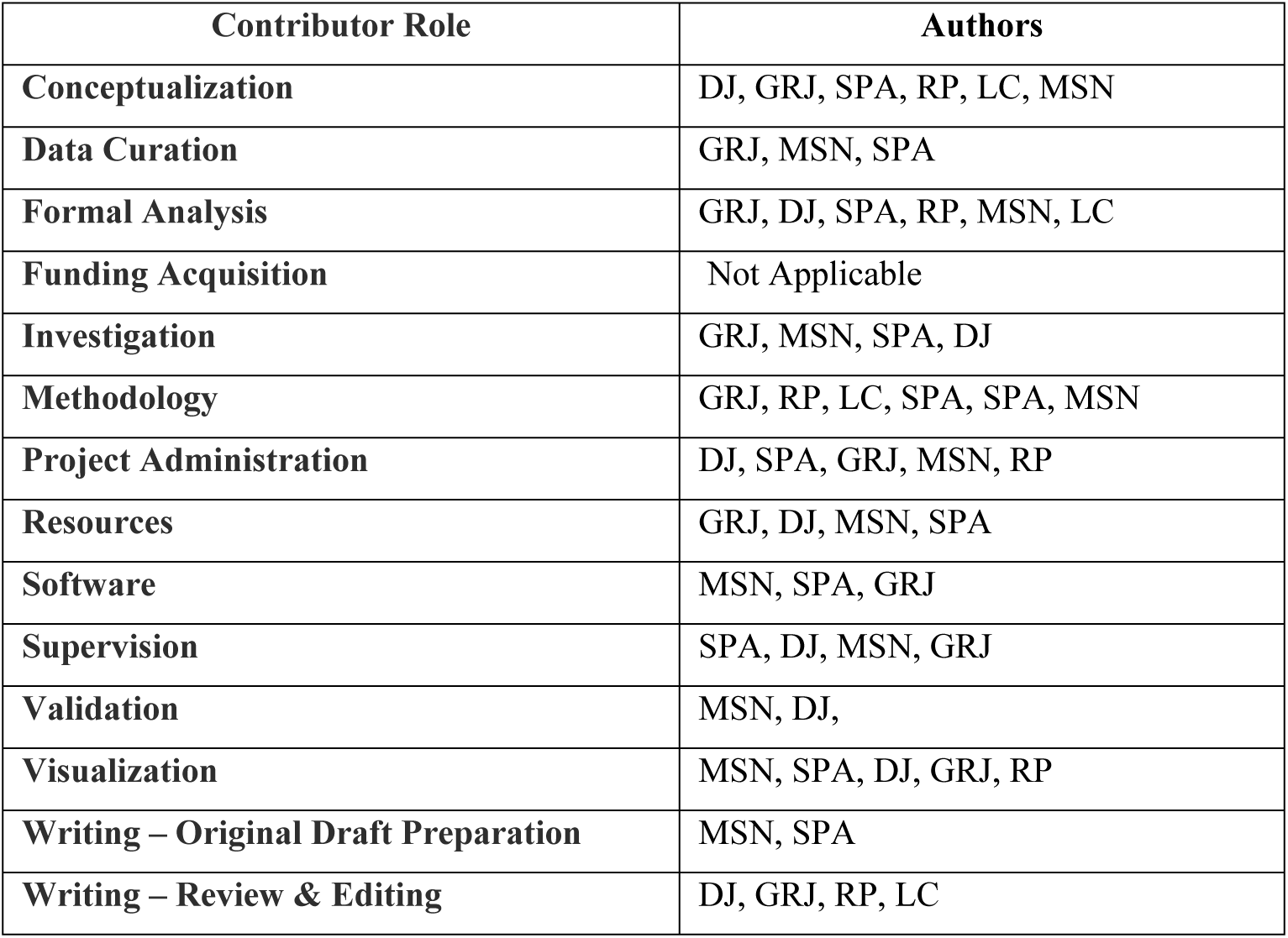

## Acknowledgment

None

## Conflict of Interest

The authors declare that they have no known competing financial interests or personal relationships that could have appeared to influence the work reported in this paper.

## Data availability statement

The synthetic population used, the AnyLogic model along with the input data are available at: https://cloud.anylogic.com/model/7cd10c0c-f1c1-4b8f-9aac-0bf37a45379a?mode=SETTINGS and https://osf.io/utmhg/?view_only=05ac26fc100645be8b1bba6557d606be.

The code developed using Python is available at: https://doi.org/10.6084/m9.figshare.19121939.

## Funding

No funding support was received for this study.

## Ethical Approval

The study has been conducted using publicly available data. No ethical approvals were sort for this study.

---

**Figure 1:**
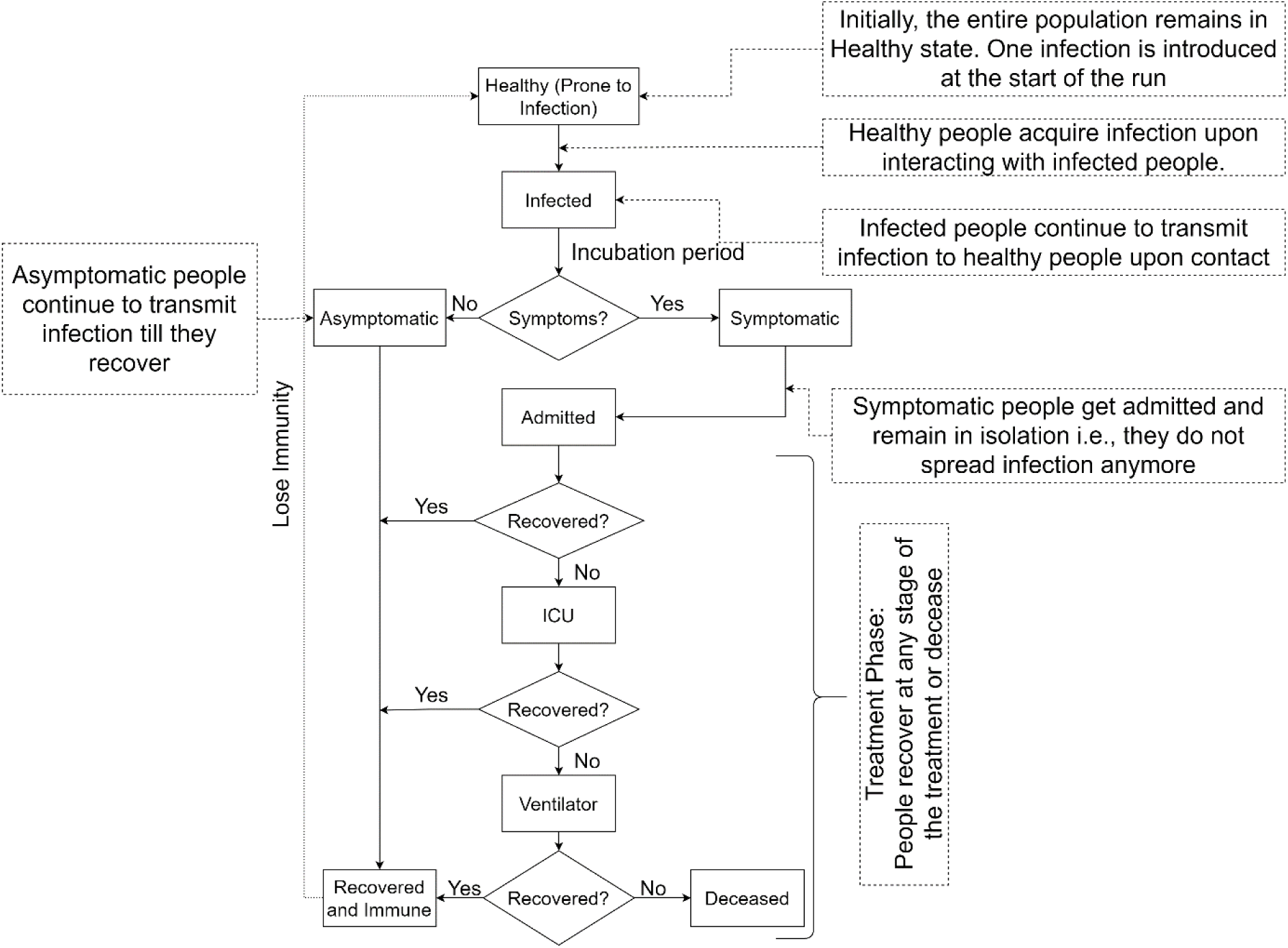
SHIVIR - Disease Model

**Figure 2:**
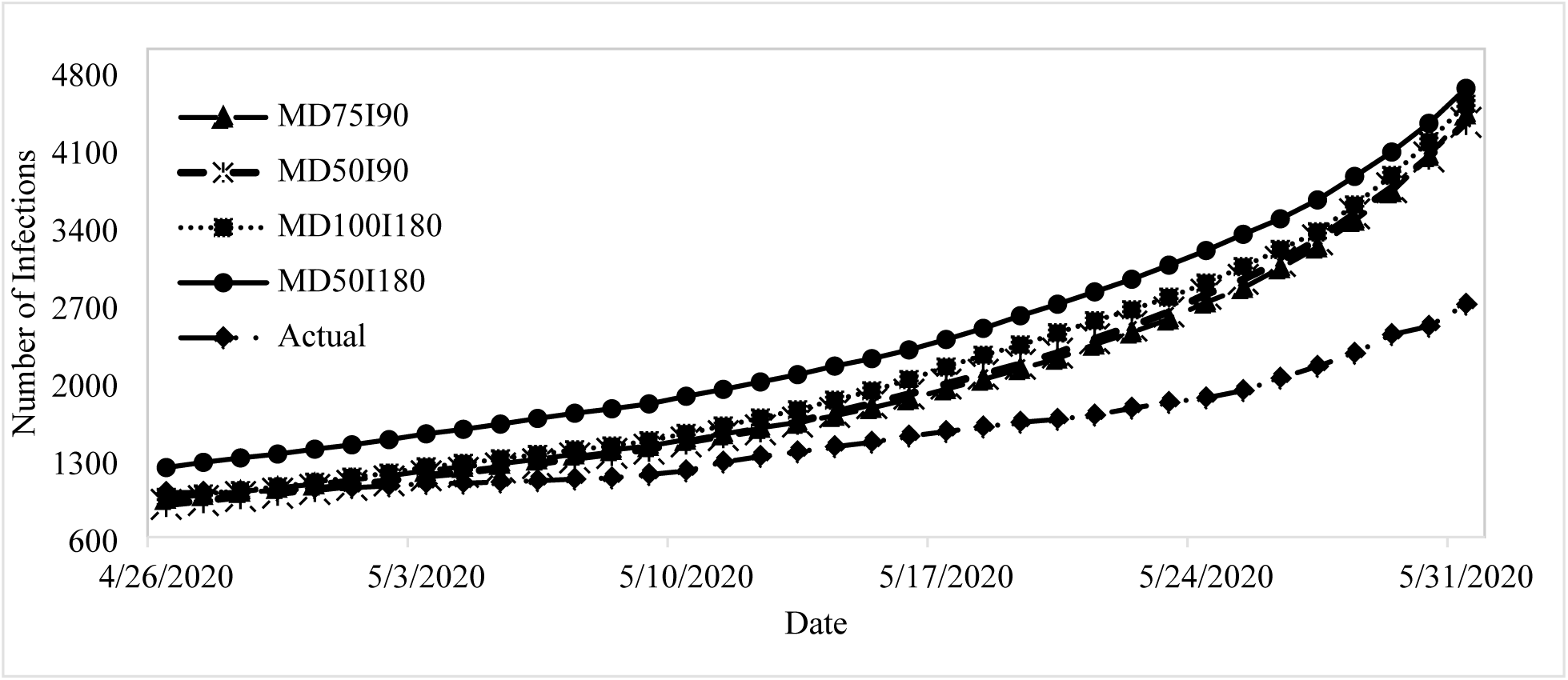
Number of Infections across scenarios

## Notes

### Competing Interest Statement

The authors have declared no competing interest.

### Funding Statement

This study did not receive any funding

